# Renin-Angiotensin-Aldosterone-System inhibitor use in patients with COVID-19 infection and prevention of serious events: a cohort study in commercially insured patients in the US

**DOI:** 10.1101/2020.07.22.20159855

**Authors:** Maria C Schneeweiss, Sandra Leonard, Andrew Weckstein, Sebastian Schneeweiss, Jeremy A Rassen

**Affiliations:** Division of Pharmacoepidemiology and Pharmacoeconomics, Department of Medicine, and Department of Dermatology Brigham and Women’s Hospital, Boston, MA; Harvard Medical School, Boston, MA; HealthVerity, Inc; Aetion, Inc

## Abstract

**Objectives:** There is lack of clarity regarding the role of angiotensin receptor blockers (ARB) or angiotensin converting enzyme inhibitors (ACEi) in interfering with the SARS-COV-2 binding on human cells and the resulting change in disease severity. We sought to assess the risk of hospitalization for COVID-19 and serious complications in current users of ARB or ACEi compared to users of dihydropyridine calcium channel blockers (dhpCCB).

**Design:** Cohort study

**Setting:** The analysis used de-identified, patient level data from HealthVerity, linking longitudinal data from US medical and pharmacy claims, which contain information on inpatient or outpatient diagnoses, procedures and medication dispensing.

**Participants:** We identified patients aged 40+ and free of chronic kidney disease (CKD) who were newly diagnosed COVID-19, between March 1, 2020 and May 30, 2020, and adherent to ACEi, ARB, or dhpCCB therapy.

**Interventions:** Current use of an ACEi, ARB, or dhpCCB.

**Main outcome measures:** We compared the 30-day risk of hospitalization for COVID-19 and serious complications.

**Results:** Of 24,708 patients identified, 7,571 were current users of an ARB, 8,484 of an ACEi, and 8,653 of a dhpCCB. The unadjusted 30-day risk of hospitalization for COVID-19 was 2.66% among ARB users, and 2.90% among ACEi users and 3.68% in dhpCCB users. In the PS-matched cohort, the risk of hospitalization among ARB users was 17% lower as compared to dhpCCB (RR=0.83; 0.68-1.00), and the risk among ACE users was 10% lower as compared to dhpCCB (RR=0.90; 0.76-1.07). When including patients with pre-existing CKD, the protective effect of ARB (RR= 0.74; 0.62-0.88) and ACEi (RR=0.84; 0.71-0.99) was more pronounced.

**Conclusions:** This cohort study showed that neither ARB nor ACEi use increase the risk of severe COVID-19 disease among those infected, and instead suggests that current use of ARB may offer a protective effect. This study found no evidence to support the discontinuation of ARB/ACEi therapy.

## Background

The spike proteins covering SARS-COV-2 bind to angiotensin-converting enzyme-2 (ACE2) receptors primarily on type II alveolar cells and cells in the heart, kidney, liver, and gastrointestinal tract, enabling the virus to inject its RNA. ^1^ It has been hypothesized that blocking the ACE2 receptor may decrease SARS-COV-2’s ability to bind to human cells, and thus reduce the severity of COVID-19 illness.^1^ However, there is uncertainty surrounding the effect of renin-angiotensin-aldosterone system (RAAS) inhibition in patients with COVID-19, with some hypothesizing that angiotensin receptor blockers (ARB) or angiotensin converting enzyme inhibitors (ACEi) use may increase the propagation of the virus. ^1-4^

Any link between ACEi or ARB use and the virus RNA’s ability to enter human cells could yield an effect, positive or negative, on the clinical severity of a COVID-19 infection, including the need for hospitalization, ventilator use, intensive care unit (ICU) stay, or death. Pre-published cohort studies by Giorgi, Mehta, and Rentsch examined the use of ACEi/ARB with respect to hospital admissions and found an increased risk; however, the contrast they made compared ACEi/ARB users to non-users and likely did not fully account for underlying factors associated with both the need for ACE or ARB treatment and with poor health outcomes.^5-7^ Khera et al. addressed some of these issues by equalizing risk through requiring comparator patients to have hypertension and to be treated with a guideline-recommended antihypertensive, and found a protective effect with ACEi use, but not ARB use, among Medicare beneficiaries.^8^ The literature lacks study findings examining the use of ACEi or ARB therapy across a broad population, and the associated risk of severe outcomes from COVID-19.^4^ As such, clinicians lack clear guidance on appropriate action to take with patients using an ACEi or ARB and diagnosed with COVID-19, in particular whether to discontinue therapy.

To that end, this study compares the risk of hospitalization for COVID-19 disease and hospitalization for serious complications of COVID-19 among prevalent users of ACEi or ARB as compared with prevalent users of dihydropyridine calcium channel blockers (dhpCCB), using recently-collected administrative claims data from the US.

## Methods

### Data source

This study used de-identified patient level healthcare data from HealthVerity covering >100 million unique patients with data from over 60 unique data sources in the US, including medical claims, pharmacy claims, lab, and electronic medical records (EMR). Data encompass all major US payer types, including commercial, Medicaid, and Medicare. For this study, we examined linked medical claims, pharmacy claims, and laboratory data containing diagnoses, procedures, prescriptions and other variables. Claims included both unadjudicated insurance claims available in near real-time time from practice management systems, billing systems, and claims clearinghouses which were then linked with adjudicated claims sourced from insurance providers and payers as well as laboratory data from hospital billing systems. Data for this study covered the period of December 1, 2018 through May 30, 2020. Claims include dates of service and diagnoses, and are recorded longitudinally, thus allowing us to establish baseline health status, COVID-19 diagnosis, and any sequalae. This study was approved by the New England IRB (#1-9757-1). De-identified patient level study data can be made available for inspection.

### Patients

Eligible patients had at least one medical encounter related to COVID-19. A COVID-19-related encounter was defined as any claim with an International Classification of Diseases, 10th Revision, Clinical Modification (ICD-10-CM) code in accordance with the Center for Disease Control’s (CDC) interim (B34.2, B97.21, B97.29, J12.81) and final (U07.1) coding guidance for confirmed cases of COVID-19^a^ or who had a positive or presumed positive COVID-19 viral lab test between March 1, 2020 and May 30, 2020 (see Appendix A for further details).

To compare the impact of ongoing ACEi or ARB therapy on severity of COVID-19 infection, we implemented a prevalent user, active comparator design where balance between treatment groups at baseline was achieved through study design and propensity score (PS) matching.^9,10^ A prevalent user design, where all study participants were using one of the study medications prior to the index date, was chosen in order to understand the effect of medications being taken at the time of COVID-19 infection, rather than the effect of initiation of therapy in response to COVID-19 diagnosis. To improve comparability among treatment groups, prevalent users of a dhpCCB were selected as the active comparator; these agents are also an anti-hypertensive treatment, like ACEi and ARB, yet due to their different mechanism of action, have no hypothesized effect on COVID-19.

We created 2 cohorts for analysis: cohort 1 included prevalent users of ARB versus dhpCCB, and cohort 2 included prevalent users of ACEi versus dhpCCB. Cohort entry was defined as the first outpatient visit with a COVID-19 diagnosis among patients with at least one prescription for ana CDC. ACEi, ARB, or dhpCCB within the 90 days before cohort entry. We excluded patients if they: (1) had no other medical or pharmacy claims activity in the 90 days prior to cohort entry, (2) qualified for more than one treatment category, (3) were of age less than 40 years on cohort entry, or (4) had a diagnosis of chronic kidney disease (CKD) within the 90 days prior to cohort entry, as CKD is a contraindication for use of an ACEi or ARB (though not for dhpCCB). We conducted a sensitivity analysis that included patients with pre-existing CKD.

### Outcomes

We compared the risk of (1) hospitalization for COVID-19 and (2) hospitalization for serious COVID-19 complications in the 30 days following cohort entry. We identified hospitalization for COVID-19 as the first inpatient visit with a diagnosis of COVID-19 (ICD-10 code: U07.1) following CDC-defined outpatient diagnosis of COVID-19. We identified hospitalization for serious COVID-19 complications as the first inpatient visit with a diagnosis of COVID-19, as above, followed on the same day or within 30 days by a diagnosis code for acute organ dysfunction or failure, acute respiratory distress syndrome (ARDS), sepsis, or respiratory intubation or mechanical ventilation (see Appendix B for a full list of codes).

Follow-up started the day after cohort entry and continued until the earliest of occurrence of outcome, death, disenrollment, 30 days, or the end of data (May 30, 2020).

### Patient characteristics

For each patient, we recorded age at cohort entry, sex, and month of cohort entry. Over the 90 days prior to cohort entry, we assessed: history of pneumonia infection requiring an office visit or hospitalization, prior use of asthma medications or anticoagulants, number of physician visits, number of unique prescription medications filled, and a range of comorbidities that may be risk factors for more severe COVID-19 disease (**Table 2**). Comorbidities included malignancy, alcohol use disorder, coagulation deficiency, HIV or AIDS, paralysis, valvular heart disease, weight loss, overweight or obesity, smoking, acute coronary syndrome, arrythmias, asthma, chronic pulmonary disease, conduction disorders, congestive heart failure, dependence on supplemental oxygen, hematologic disorders, hypertension, inflammatory heart disease, ischemic heart disease, liver disease, myocardial infarction, stroke, pulmonary circulation disorders, rheumatic disease, and systemic lupus erythematosus.

### Statistical analysis

We tabulated baseline patient characteristics and assessed the 30-day risk of the outcomes of interest with 95% confidence intervals for each exposure group.

To account for potential confounding by measured patient characteristics, we used propensity score (PS)-matching to achieve balance of covariates between the treatment groups.^11,12^ While a prevalent user analysis can lead to bias from adjusting for intermediate variables,^13^ in this case both the drug exposure and the patient characteristics were observed before cohort entry, and thus are unlikely to cause bias. The PS, defined as the probability that a patient was using either ACEi/ARB versus dhpCCB, conditioning on all the confounders listed above, was estimated with a multivariable logistic regression model. PS-matching was performed using 1:1 nearest neighbor matching with a maximum matching caliper of 0.02 on the PS scale.^14^ To quantify balance achieved at baseline, we computed standardized differences in the covariates between the two groups after PS-matching.^15^

We calculated relative risks (RR) and 95% confidence intervals (CIs) of the outcomes associated with drug use using logistic regression. All analyses were conducted using the Aetion Evidence Platform v3.19 (including R version 4.0.2), which has been validated and has successfully predicted randomized control trial findings.^16-18^ All analyses from the raw patient-level data are recorded with audit trails and can be reviewed on request.

## Results

We identified 24,708 patients aged 40 years or older who had an outpatient diagnosis of COVID-19 and met the inclusion criteria (**Table 1**). At the start of follow-up, cohort 1 had 15,829 prevalent users of either an ARB (n=7,571) or dhpCCB (n=8,258), while cohort 2 had 17,137 prevalent users of either an ACEi (n=8,484) or dhpCCB (n=8,653) (**Tables 2, 3**). After applying 1:1 PS-matching on all baseline characteristics, cohort 1 had 6,710 pairs of patients using ARB vs. dhpCCB and cohort 2 had 7,334 pairs of patients using ACEi vs. dhpCCB. In the PS-matched cohort, all 60 baseline characteristics were well-balanced between treatment groups (**Tables 2, 3**) with all absolute standardized differences between treatment groups below the recognized threshold of 0.1.^19^

**Table 1:**
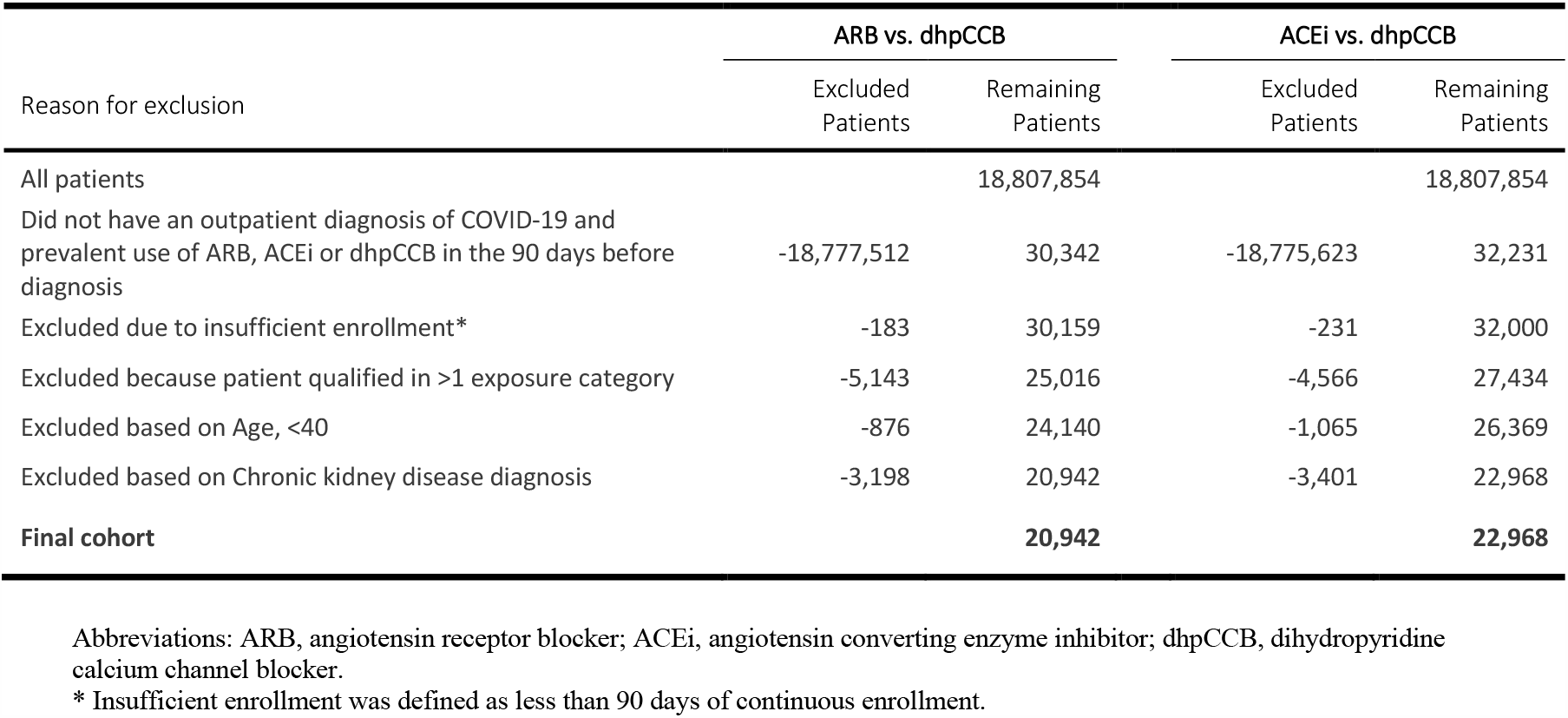
Application of inclusion/exclusion criteria.

**Table 2:** Baseline patient characteristics, comparing users of ARB vs. dhpCCB*.

| Variable | Unmatched patients |  | After 1:1 propensity-score matching |  | Standardized difference |
| --- | --- | --- | --- | --- | --- |
|  | ARB | dhpCCB | ARB | dhpCCB |  |
| Number of patients | 7,571 | 8,258 | 6,710 | 6,710 |  |
| Month/year of cohort entry date |  |  |  |  | 0.015 |
| ...March 2020; n (%) | 1,591 (21.0%) | 1,441 (17.4%) | 1,291 (19.2%) | 1,315 (19.6%) |  |
| ...April 2020; n (%) | 5,029 (66.4%) | 5,628 (68.2%) | 4,535 (67.6%) | 4,487 (66.9%) |  |
| ...May 2020; n (%) | 951 (12.6%) | 1,189 (14.4%) | 884 (13.2%) | 908 (13.5%) |  |
| Age; mean (sd) | 66.80 (13.35) | 68.87 (13.93) | 67.52 (13.33) | 67.48 (13.78) | 0.003 |
| Female; n (%) | 4,148 (54.8%) | 4,283 (51.9%) | 3,590 (53.5%) | 3,596 (53.6%) | 0.014 |
| Alcohol abuse; n (%) | 47 (0.6%) | 114 (1.4%) | 45 (0.7%) | 45 (0.7%) | 0.000 |
| Coagulation deficiency; n (%) | 78 (1.0%) | 95 (1.2%) | 66 (1.0%) | 62 (0.9%) | 0.006 |
| Depression; n (%) | 449 (5.9%) | 707 (8.6%) | 419 (6.2%) | 396 (5.9%) | 0.014 |
| Drug abuse; n (%) | 45 (0.6%) | 103 (1.2%) | 43 (0.6%) | 48 (0.7%) | 0.009 |
| Fluid and electrolyte disorders; n (%) | 23 (0.3%) | 25 (0.3%) | 19 (0.3%) | 17 (0.3%) | 0.006 |
| HIV and AIDS; n (%) | 24 (0.3%) | 44 (0.5%) | 24 (0.4%) | 29 (0.4%) | 0.012 |
| Lymphoma; n (%) | 29 (0.4%) | 36 (0.4%) | 29 (0.4%) | 26 (0.4%) | 0.007 |
| Metastatic cancer; n (%) | 44 (0.6%) | 62 (0.8%) | 41 (0.6%) | 40 (0.6%) | 0.002 |
| Paralysis; n (%) | 49 (0.6%) | 112 (1.4%) | 48 (0.7%) | 51 (0.8%) | 0.005 |
| Psychoses; n (%) | 103 (1.4%) | 246 (3.0%) | 103 (1.5%) | 87 (1.3%) | 0.020 |
| Solid tumor without metastasis; n (%) | 238 (3.1%) | 278 (3.4%) | 217 (3.2%) | 216 (3.2%) | 0.001 |
| Valvular disease; n (%) | 202 (2.7%) | 164 (2.0%) | 153 (2.3%) | 136 (2.0%) | 0.017 |
| Weight loss; n (%) | 80 (1.1%) | 159 (1.9%) | 78 (1.2%) | 76 (1.1%) | 0.003 |
| Smoking; n (%) | 186 (2.5%) | 239 (2.9%) | 172 (2.6%) | 163 (2.4%) | 0.009 |
| Pregnancy; n (%) | 4 (0.1%) | 1 (0.0%) | 2 (0.0%) | 1 (0.0%) | 0.010 |
| Acute coronary syndrome; n (%) | 96 (1.3%) | 89 (1.1%) | 80 (1.2%) | 74 (1.1%) | 0.008 |
| Arrhythmias or cardiac dysrhythmias; n (%) | 672 (8.9%) | 683 (8.3%) | 557 (8.3%) | 532 (7.9%) | 0.014 |
| Asthma; n (%) | 346 (4.6%) | 303 (3.7%) | 248 (3.7%) | 251 (3.7%) | 0.002 |
| Cerebrovascular diseases; n (%) | 342 (4.5%) | 632 (7.7%) | 324 (4.8%) | 326 (4.9%) | 0.001 |
| Chronic kidney disease a; n (%) | 0 (0.0%) a | 0 (0.0%) a | 0 (0.0%) a | 0 (0.0%) a | - |
| Chronic pulmonary disease; n (%) | 781 (10.3%) | 897 (10.9%) | 638 (9.5%) | 645 (9.6%) | 0.004 |
| Cirrhosis; n (%) | 25 (0.3%) | 27 (0.3%) | 19 (0.3%) | 21 (0.3%) | 0.005 |
| Conduction disorders; n (%) | 114 (1.5%) | 120 (1.5%) | 97 (1.4%) | 83 (1.2%) | 0.018 |
| Congestive heart failure; n (%) | 584 (7.7%) | 508 (6.2%) | 448 (6.7%) | 428 (6.4%) | 0.012 |
| Cystic fibrosis; n (%) | 0 (0.0%) | 0 (0.0%) | 0 (0.0%) | 0 (0.0%) | - |
| Dependence on supplemental oxygen; n (%) | 99 (1.3%) | 136 (1.6%) | 91 (1.4%) | 92 (1.4%) | 0.001 |
| End-stage renal disease a; n (%) | 0 (0.0%) a | 0 (0.0%) a | 0 (0.0%) a | 0 (0.0%) a | - |
| Hematologic disorders; n (%) | 92 (1.2%) | 89 (1.1%) | 73 (1.1%) | 66 (1.0%) | 0.010 |
| Hyperlipidemia; n (%) | 1,514 (20.0%) | 1,522 (18.4%) | 1,226 (18.3%) | 1,202 (17.9%) | 0.009 |
| Hypertension; n (%) | 2,702 (35.7%) | 3,180 (38.5%) | 2,329 (34.7%) | 2,298 (34.2%) | 0.010 |
| Inflammatory heart disorders (pericarditis, endocarditis, myocarditis); n (%) | 8 (0.1%) | 10 (0.1%) | 7 (0.1%) | 5 (0.1%) | 0.010 |
| Ischemic heart disease; n (%) | 691 (9.1%) | 670 (8.1%) | 564 (8.4%) | 529 (7.9%) | 0.019 |
| Liver disease, mild; n (%) | 119 (1.6%) | 140 (1.7%) | 103 (1.5%) | 103 (1.5%) | 0.000 |
| Liver disease, moderate or severe; n (%) | 9 (0.1%) | 11 (0.1%) | 7 (0.1%) | 7 (0.1%) | 0.000 |
| Myocardial Infarction; n (%) | 128 (1.7%) | 130 (1.6%) | 108 (1.6%) | 106 (1.6%) | 0.002 |
| Overweight or obesity; n (%) | 966 (12.8%) | 830 (10.1%) | 741 (11.0%) | 735 (11.0%) | 0.003 |
| Obstructive sleep apnea; n (%) | 351 (4.6%) | 274 (3.3%) | 258 (3.8%) | 249 (3.7%) | 0.007 |
| Peptic ulcer disease; n (%) | 24 (0.3%) | 47 (0.6%) | 23 (0.3%) | 14 (0.2%) | 0.026 |
| Peripheral vascular disease; n (%) | 386 (5.1%) | 472 (5.7%) | 337 (5.0%) | 327 (4.9%) | 0.007 |
| Potassium or magnesium imbalance; n (%) | 10 (0.1%) | 9 (0.1%) | 5 (0.1%) | 7 (0.1%) | 0.010 |
| Psoriasis; n (%) | 31 (0.4%) | 21 (0.3%) | 25 (0.4%) | 17 (0.3%) | 0.021 |
| Pulmonary circulation disorders; n (%) | 97 (1.3%) | 113 (1.4%) | 81 (1.2%) | 82 (1.2%) | 0.001 |
| Renal impairment; n (%) | 267 (3.5%) | 418 (5.1%) | 251 (3.7%) | 256 (3.8%) | 0.004 |
| Rheumatic disease; n (%) | 130 (1.7%) | 122 (1.5%) | 106 (1.6%) | 101 (1.5%) | 0.006 |
| Stroke (ischemic, hemorrhagic); n (%) | 148 (2.0%) | 299 (3.6%) | 144 (2.1%) | 148 (2.2%) | 0.004 |
| Systemic lupus erythematosus; n (%) | 14 (0.2%) | 17 (0.2%) | 12 (0.2%) | 9 (0.1%) | 0.011 |
| Type-2 Diabetes; n (%) | 1,562 (20.6%) | 1,608 (19.5%) | 1,272 (19.0%) | 1,251 (18.6%) | 0.008 |
| All cause pneumonia; n (%) | 651 (8.6%) | 834 (10.1%) | 598 (8.9%) | 589 (8.8%) | 0.005 |
| Use of any anticoagulant; n (%) | 797 (10.5%) | 808 (9.8%) | 657 (9.8%) | 664 (9.9%) | 0.004 |
| Use of any novel oral anticoagulant; n (%) | 631 (8.3%) | 622 (7.5%) | 529 (7.9%) | 533 (7.9%) | 0.002 |
| Use of warfarin; n (%) | 154 (2.0%) | 133 (1.6%) | 113 (1.7%) | 118 (1.8%) | 0.006 |
| Use of any anti-platelet medication; n (%) | 1,075 (14.2%) | 1,356 (16.4%) | 998 (14.9%) | 975 (14.5%) | 0.010 |
| Use of corticosteroid, inhaled; n (%) | 749 (9.9%) | 691 (8.4%) | 589 (8.8%) | 588 (8.8%) | 0.001 |
| Use of any asthma medication; n (%) | 2,372 (31.3%) | 2,374 (28.7%) | 2,002 (29.8%) | 1,999 (29.8%) | 0.001 |
| Any inpatient medical claim; n (%) | 1,452 (19.2%) | 2,148 (26.0%) | 1,354 (20.2%) | 1,305 (19.4%) | 0.018 |
| Coronary care unit visit; n (%) | 20 (0.3%) | 26 (0.3%) | 20 (0.3%) | 23 (0.3%) | 0.008 |
| Intensive care unit revenue codes; n (%) | 64 (0.8%) | 68 (0.8%) | 50 (0.7%) | 48 (0.7%) | 0.004 |
| No. of any physician visit; mean (sd) | 1.18 (2.50) | 0.92 (2.46) | 1.04 (2.14) | 1.01 (2.65) | 0.010 |
| No. of pharmacy dispensings; mean (sd) | 6.23 (6.18) | 6.61 (6.86) | 6.25 (6.27) | 6.18 (6.39) | 0.011 |

Abbreviations: ARB, angiotensin receptor blocker; ACEi, angiotensin converting enzyme inhibitor; dhpCCB, dihydropyridine calcium channel blocker; sd, standard deviation; No., number.
\* Patients were ongoing users of either ARB or dhpCCB in the 90 days before cohort entry (documented COVID-19 infection); all patients characteristics were assessed during the 90 days before cohort entry.
<sup>a</sup> The baseline prevalence of chronic kidney disease and end stage renal disease was 0% as it was a study exclusion criterion.

**Table 3:** Baseline patient characteristics, comparing users of ACEi vs. dhpCCB*.

| Variable | Unmatched patients |  | After 1:1 propensity-score matching |  | Standardized difference |
| --- | --- | --- | --- | --- | --- |
|  | ACEi | dhpCCB | ACEi | dhpCCB |  |
| Number of patients | 8,484 | 8,653 | 7,334 | 7,334 |  |
| Month/year of cohort entry date |  |  |  |  | 0.009 |
| ...March 2020; n (%) | 1,527 (18.0%) | 1,592 (18.4%) | 1,306 (17.8%) | 1,330 (18.1%) |  |
| ...April 2020; n (%) | 5,712 (67.3%) | 5,876 (67.9%) | 4,976 (67.8%) | 4,965 (67.7%) |  |
| ...May 2020; n (%) | 1,245 (14.7%) | 1,185 (13.7%) | 1,052 (14.3%) | 1,039 (14.2%) |  |
| Age; mean (sd) | 66.06 (13.19) | 68.90 (13.91) | 66.97 (13.17) | 67.13 (13.48) | 0.012 |
| Female; n (%) | 3,990 (47.0%) | 4,706 (54.4%) | 3,695 (50.4%) | 3,680 (50.2%) | 0.004 |
| Alcohol abuse; n (%) | 109 (1.3%) | 106 (1.2%) | 93 (1.3%) | 87 (1.2%) | 0.007 |
| Coagulation deficiency; n (%) | 102 (1.2%) | 88 (1.0%) | 67 (0.9%) | 75 (1.0%) | 0.011 |
| Depression; n (%) | 649 (7.6%) | 645 (7.5%) | 530 (7.2%) | 506 (6.9%) | 0.013 |
| Drug abuse; n (%) | 104 (1.2%) | 87 (1.0%) | 87 (1.2%) | 79 (1.1%) | 0.010 |
| Fluid and electrolyte disorders; n (%) | 22 (0.3%) | 17 (0.2%) | 14 (0.2%) | 14 (0.2%) | 0.000 |
| HIV and AIDS; n (%) | 47 (0.6%) | 37 (0.4%) | 37 (0.5%) | 36 (0.5%) | 0.002 |
| Lymphoma; n (%) | 31 (0.4%) | 47 (0.5%) | 27 (0.4%) | 27 (0.4%) | 0.000 |
| Metastatic cancer; n (%) | 42 (0.5%) | 66 (0.8%) | 39 (0.5%) | 32 (0.4%) | 0.014 |
| Paralysis; n (%) | 85 (1.0%) | 105 (1.2%) | 76 (1.0%) | 83 (1.1%) | 0.009 |
| Psychoses; n (%) | 222 (2.6%) | 195 (2.3%) | 166 (2.3%) | 167 (2.3%) | 0.001 |
| Solid tumor without metastasis; n (%) | 257 (3.0%) | 287 (3.3%) | 221 (3.0%) | 211 (2.9%) | 0.008 |
| Valvular disease; n (%) | 213 (2.5%) | 186 (2.1%) | 145 (2.0%) | 156 (2.1%) | 0.011 |
| Weight loss; n (%) | 121 (1.4%) | 152 (1.8%) | 106 (1.4%) | 100 (1.4%) | 0.007 |
| Smoking; n (%) | 275 (3.2%) | 246 (2.8%) | 202 (2.8%) | 208 (2.8%) | 0.005 |
| Pregnancy; n (%) | 1 (0.0%) | 0 (0.0%) | 0 (0.0%) | 0 (0.0%) | - |
| Acute coronary syndrome; n (%) | 108 (1.3%) | 88 (1.0%) | 77 (1.0%) | 76 (1.0%) | 0.001 |
| Arrhythmias or cardiac dysrhythmias; n (%) | 803 (9.5%) | 693 (8.0%) | 581 (7.9%) | 593 (8.1%) | 0.006 |
| Asthma; n (%) | 308 (3.6%) | 339 (3.9%) | 267 (3.6%) | 265 (3.6%) | 0.001 |
| Cerebrovascular diseases; n (%) | 482 (5.7%) | 589 (6.8%) | 434 (5.9%) | 416 (5.7%) | 0.011 |
| Chronic kidney disease a; n (%) | 0 (0.0%) a | 0 (0.0%) a | 0 (0.0%) a | 0 (0.0%) a | - |
| Chronic pulmonary disease; n (%) | 884 (10.4%) | 928 (10.7%) | 748 (10.2%) | 750 (10.2%) | 0.001 |
| Cirrhosis; n (%) | 43 (0.5%) | 32 (0.4%) | 31 (0.4%) | 31 (0.4%) | 0.000 |
| Conduction disorders; n (%) | 117 (1.4%) | 118 (1.4%) | 88 (1.2%) | 86 (1.2%) | 0.003 |
| Congestive heart failure; n (%) | 655 (7.7%) | 523 (6.0%) | 452 (6.2%) | 464 (6.3%) | 0.007 |
| Cystic fibrosis; n (%) | 0 (0.0%) | 0 (0.0%) | 0 (0.0%) | 0 (0.0%) | - |
| Dependence on supplemental oxygen; n (%) | 132 (1.6%) | 150 (1.7%) | 111 (1.5%) | 111 (1.5%) | 0.000 |
| End-stage renal disease a; n (%) | 0 (0.0%) a | 0 (0.0%) a | 0 (0.0%) a | 0 (0.0%) a | - |
| Hematologic disorders; n (%) | 119 (1.4%) | 103 (1.2%) | 80 (1.1%) | 90 (1.2%) | 0.013 |
| Hyperlipidemia; n (%) | 1,732 (20.4%) | 1,595 (18.4%) | 1,356 (18.5%) | 1,378 (18.8%) | 0.008 |
| Hypertension; n (%) | 3,003 (35.4%) | 3,277 (37.9%) | 2,561 (34.9%) | 2,575 (35.1%) | 0.004 |
| Inflammatory heart disorders (pericarditis, endocarditis, myocarditis); n (%) | 7 (0.1%) | 10 (0.1%) | 7 (0.1%) | 8 (0.1%) | 0.004 |
| Ischemic heart disease; n (%) | 760 (9.0%) | 679 (7.8%) | 560 (7.6%) | 573 (7.8%) | 0.007 |
| Liver disease, mild; n (%) | 171 (2.0%) | 142 (1.6%) | 131 (1.8%) | 121 (1.6%) | 0.010 |
| Liver disease, moderate or severe; n (%) | 18 (0.2%) | 12 (0.1%) | 14 (0.2%) | 12 (0.2%) | 0.006 |
| Myocardial Infarction; n (%) | 171 (2.0%) | 121 (1.4%) | 106 (1.4%) | 109 (1.5%) | 0.003 |
| Overweight or obesity; n (%) | 1,022 (12.0%) | 912 (10.5%) | 794 (10.8%) | 810 (11.0%) | 0.007 |
| Obstructive sleep apnea; n (%) | 323 (3.8%) | 344 (4.0%) | 273 (3.7%) | 295 (4.0%) | 0.016 |
| Peptic ulcer disease; n (%) | 24 (0.3%) | 40 (0.5%) | 22 (0.3%) | 23 (0.3%) | 0.002 |
| Peripheral vascular disease; n (%) | 479 (5.6%) | 446 (5.2%) | 365 (5.0%) | 370 (5.0%) | 0.003 |
| Potassium or magnesium imbalance; n (%) | 3 (0.0%) | 7 (0.1%) | 3 (0.0%) | 3 (0.0%) | 0.000 |
| Psoriasis; n (%) | 21 (0.2%) | 25 (0.3%) | 21 (0.3%) | 19 (0.3%) | 0.005 |
| Pulmonary circulation disorders; n (%) | 102 (1.2%) | 123 (1.4%) | 84 (1.1%) | 84 (1.1%) | 0.000 |
| Renal impairment; n (%) | 308 (3.6%) | 417 (4.8%) | 285 (3.9%) | 283 (3.9%) | 0.001 |
| Rheumatic disease; n (%) | 93 (1.1%) | 141 (1.6%) | 82 (1.1%) | 89 (1.2%) | 0.009 |
| Stroke (ischemic, hemorrhagic); n (%) | 228 (2.7%) | 278 (3.2%) | 207 (2.8%) | 207 (2.8%) | 0.000 |
| Systemic lupus erythematosus; n (%) | 20 (0.2%) | 25 (0.3%) | 15 (0.2%) | 19 (0.3%) | 0.011 |
| Type-2 Diabetes; n (%) | 2,029 (23.9%) | 1,603 (18.5%) | 1,446 (19.7%) | 1,486 (20.3%) | 0.014 |
| All cause pneumonia; n (%) | 745 (8.8%) | 869 (10.0%) | 650 (8.9%) | 668 (9.1%) | 0.009 |
| Use of any anticoagulant; n (%) | 885 (10.4%) | 813 (9.4%) | 688 (9.4%) | 697 (9.5%) | 0.004 |
| Use of any novel oral anticoagulant; n (%) | 645 (7.6%) | 614 (7.1%) | 524 (7.1%) | 534 (7.3%) | 0.005 |
| Use of warfarin; n (%) | 217 (2.6%) | 147 (1.7%) | 134 (1.8%) | 139 (1.9%) | 0.005 |
| Use of any anti-platelet medication; n (%) | 1,259 (14.8%) | 1,406 (16.2%) | 1,100 (15.0%) | 1,103 (15.0%) | 0.001 |
| Use of corticosteroid, inhaled; n (%) | 667 (7.9%) | 786 (9.1%) | 595 (8.1%) | 638 (8.7%) | 0.021 |
| Use of any asthma medication; n (%) | 2,260 (26.6%) | 2,636 (30.5%) | 2,037 (27.8%) | 2,096 (28.6%) | 0.018 |
| Any inpatient medical claim; n (%) | 1,879 (22.1%) | 2,068 (23.9%) | 1,593 (21.7%) | 1,613 (22.0%) | 0.007 |
| Coronary care unit visit; n (%) | 25 (0.3%) | 21 (0.2%) | 19 (0.3%) | 15 (0.2%) | 0.011 |
| Intensive care unit revenue codes; n (%) | 67 (0.8%) | 65 (0.8%) | 49 (0.7%) | 50 (0.7%) | 0.002 |
| No. of any physician visit; mean (sd) | 1.06 (2.49) | 0.95 (2.30) | 0.94 (2.12) | 0.96 (2.32) | 0.007 |
| No. of pharmacy dispensings; mean (sd) | 6.17 (6.31) | 6.54 (6.66) | 6.18 (6.36) | 6.22 (6.44) | 0.007 |

Abbreviations: ARB, angiotensin receptor blocker; ACEi, angiotensin converting enzyme inhibitor; dhpCCB, dihydropyridine calcium channel blocker; sd, standard deviation; No., number.
\* Patients were ongoing users of either ACEi or dhpCCB in the 90 days before cohort entry (documented COVID-19 infection); all patients characteristics were assessed during the 90 days before cohort entry.
<sup>a</sup> The baseline prevalence of chronic kidney disease and end stage renal disease was 0% as it was a study exclusion criterion.

In the PS-matched cohort, the 30-day risk of hospitalization for COVID-19 was 2.85% in ARB users (vs. 3.44% in dhpCCB users) and 3.12% in ACEi users (vs. 3.46% in dhpCCB users) (**Table 4**). After PS-matching we found that ARB users had a 17% decrease in the risk of hospitalization for COVID-19 (RR=0.83; 95% CI, 0.68-1.00) as compared to dhpCCB, while ACEi users had a 10% decrease in risk for hospitalization for COVID-19 (RR=0.90; 0.76-1.07) as compared to dhpCCB. Point estimates suggested protective effects for several serious complications, though confidence intervals generally included the null: acute organ dysfunction or failure (RR=0.88 [95% CI: 0.65-1.20] for ARB and 1.00 [0.76-1.32] for ACEi); ARDS (RR=0.86 [0.63-1.16] for ARB and 0.99 [0.75-1.31] for ACEi); sepsis (RR=0.71 [0.49-1.03] for ARB and 1.06 [0.76-1.48] for ACEi); and for respiratory intubation or mechanical ventilation (RR=0.77 [0.59-1.00] for ARB and 0.99 [0.78-1.26] for ACEi) (**Table 4**).

**Table 4:** 30-day risk and relative risk of hospitalization for COVID-19 infection among users of ARB vs. dhpCCB or ACEi vs. dhpCCB.

| Outcome | Comparison | Unmatched patients |  |  |  |  | 1:1 propensity score matched patients |  |  |  |  |
| --- | --- | --- | --- | --- | --- | --- | --- | --- | --- | --- | --- |
|  |  | N of patients | N of events | 30-day risk | Risk Ratio (95% CI) | Risk difference (95% CI) | N of patients | N of events | 30-day risk | Risk Ratio (95% CI) | Risk difference (95% CI) |
| **Hospitalization for COVID-19** | **ARB** | 7,571 | 201 | 2.66% | 0.72 (0.61, 0.86) | -1.02% (-1.58, -0.47) | 6,710 | 191 | 2.85% | 0.83 (0.68, 1.00) | -0.59% (-1.20, 0.01) |
|  | dhpCCB | 8,258 | 304 | 3.68% | referent | referent | 6,710 | 231 | 3.44% | referent | referent |
|  | **ACEi** | 8,484 | 246 | 2.90% | 0.79 (0.67, 0.93) | -0.76% (-1.31, -0.22) | 7,334 | 229 | 3.12% | 0.90 (0.76, 1.07) | -0.34% (-0.93, 0.25) |
|  | dhpCCB | 8,653 | 317 | 3.66% | referent | referent | 7,334 | 254 | 3.46% | referent | referent |
| **Hospitalization with acute organ dysfunction<sup>b</sup>** | **ARB** | 7,571 | 85 | 1.12% | 0.81 (0.62, 1.08) | -0.26% (-0.62, 0.10) | 6,710 | 76 | 1.13% | 0.88 (0.65, 1.20) | -0.15% (-0.53, 0.24) |
|  | dhpCCB | 8,258 | 114 | 1.38% | referent | referent | 6,710 | 86 | 1.28% | referent | referent |
|  | **ACEi** | 8,484 | 104 | 1.23% | 0.94 (0.72, 1.22) | -0.08% (-0.43, 0.27) | 7,334 | 97 | 1.32% | 1.00 (0.76, 1.32) | 0.00 (-0.37, 0.37) |
|  | dhpCCB | 8,653 | 113 | 1.31% | referent | referent | 7,334 | 97 | 1.32% | referent | referent |
| **Hospitalization with acute respiratory distress syndrome** | **ARB** | 7,571 | 87 | 1.15% | 0.78 (0.60, 1.03) | -0.32% (-0.68, 0.05) | 6,710 | 78 | 1.16% | 0.86 (0.63, 1.16) | -0.20% (-0.59, 0.20) |
|  | dhpCCB | 8,258 | 121 | 1.47% | referent | referent | 6,710 | 91 | 1.36% | referent | referent |
|  | **ACEi** | 8,484 | 102 | 1.20% | 0.90 (0.69, 1.18) | -0.13% (-0.47, 0.22) | 7,334 | 97 | 1.32% | 0.99 (0.75, 1.31) | -0.01% (-0.40, 0.37) |
|  | dhpCCB | 8,653 | 115 | 1.33% | referent | referent | 7,334 | 98 | 1.33% | referent | referent |
| **Hospitalization with sepsis** | **ARB** | 7,571 | 54 | 0.71% | 0.69 (0.49, 0.97) | -0.32% (-0.62, -0.02) | 6,710 | 46 | 0.69% | 0.71 (0.49, 1.03) | -0.28% (-0.60, 0.04) |
|  | dhpCCB | 8,258 | 85 | 1.03% | referent | referent | 6,710 | 65 | 0.97% | referent | referent |
|  | **ACEi** | 8,484 | 76 | 0.89% | 0.92 (0.68, 1.26) | -0.08% (-0.38, 0.23) | 7,334 | 71 | 0.97% | 1.06 (0.76, 1.48) | 0.06% (-0.27, 0.38) |
|  | dhpCCB | 8,653 | 84 | 0.97% | referent | referent | 7,334 | 67 | 0.91% | referent | referent |
| **Hospitalization with respiratory intubation or mechanical ventilation** | **ARB** | 7,571 | 113 | 1.49% | 0.87 (0.68, 1.11) | -0.23% (-0.63, 0.18) | 6,710 | 92 | 1.37% | 0.77 (0.59, 1.00) | -0.42% (-0.85, 0.02) |
|  | dhpCCB | 8,258 | 142 | 1.72% | referent | referent | 6,710 | 120 | 1.79% | referent | referent |
|  | **ACEi** | 8,484 | 158 | 1.86% | 0.99 (0.80, 1.23) | -0.02% (-0.44, 0.39) | 7,334 | 131 | 1.79% | 0.99 (0.78, 1.26) | -0.01% (-0.46, 0.43) |
|  | dhpCCB | 8,653 | 163 | 1.88% | referent | referent | 7,334 | 132 | 1.80% | referent | referent |

Abbreviations: ARB, angiotensin receptor blocker; ACEi, angiotensin converting enzyme inhibitor; dhpCCB, dihydropyridine calcium channel blocker; N, number; Pts, patients; 95% CI, 95% confidence interval.
<sup>a</sup> The baseline prevalence of chronic kidney disease and end stage renal disease was 0% as it was a study exclusion criterion; <sup>b</sup> Acute organ dysfunction includes: Critical illness polyneuropathy, Acute kidney failure unspecified, Metabolic encephalopathy, Acute respiratory failure, Acute respiratory failure with hypoxia, Acute and subacute hepatic failure with coma, Acute kidney failure, Acute kidney failure with acute cortical necrosis, Disseminated intravascular coagulation, Acute and subacute hepatic failure, Acute and subacute hepatic failure without coma, Acute kidney failure with medullary necrosis, Acute respiratory failure, unspecified whether with hypoxia or hypercapnia, Acute respiratory failure with hypercapnia, Other acute kidney failure, Critical illness myopathy, Acute kidney failure with tubular necrosis

In our sensitivity analysis including patients with pre-existing CKD in the 90 days before outpatient COVID-19 diagnosis, the relative risk of hospitalization for COVID-19 decreased to 0.74 (0.62-0.88) for ARB and 0.84 (0.71-0.99) for ACEi.

## Discussion

In this population-representative study of patients aged 40 and older with an outpatient COVID-19 diagnosis, we observed a 17% decrease in the 30-day risk of hospitalization for COVID-19 among ARB users compared to dhpCCB users, and a 10% decrease in risk of hospitalization for COVID-19 among ACEi users compared to dhpCCB users. We also observed a numerically decreased 30-day risk of respiratory intubation and mechanical ventilation after hospitalization with COVID-19 among ARB users compared to dhpCCB users. There was no meaningful difference in the risk of acute organ dysfunction, ARDS, or sepsis among RAAS inhibitors vs. dhpCCBs.

Our findings suggest neither a need to discontinue ACEi/ARB therapy among patients using those medications at the time of COVID-19 diagnosis, nor a clear case for use of ACEi/ARB therapy as prophylaxis among COVID-19-infected patients.

There were several notable strengths of this study. First, we used a large population-representative cohort with over 20,000 COVID-19 adults 40+ receiving anti-hypertensive treatment with RAAS inhibitors or dhpCCB from established data sources. Second, the outcome of interest was severe enough to avoid differential surveillance bias since all serious infections that require hospitalization are captured in claims data, and the ICD codes used to identify them have been shown to be of high accuracy.^20^ Third, we implemented robust adjustment for potential confounders using active comparator study design combined with 1:1 propensity score matching.^21,22^ Fourth, by requiring comparator patients also be treated with an antihypertensive medication, the confounder-adjusted, dhpCCB control group provided an appropriate reference value for hospitalization in patients aged 40 or older with COVID-19 infection.

This population-based study applying a causal study design concludes that neither ARB nor ACEi use increase the risk of severe COVID-disease among those infected, and rather suggest that prevalent use of these medications – ARB in particular -- may offer a protective effect. Indeed, our findings suggest a small decrease in risk of hospitalization for COVID-19 between adult patients with an outpatient COVID-19 diagnosis who were using RAAS inhibitors versus those treated with dhpCCB. These findings are reassuring as they suggest that RAAS inhibitors should not be discontinued among patients with COVID-19 infection.

## Data Availability

De-identified patient level study data can be made available for inspection. All analyses from the raw patient-level data are recorded with audit trails and can be reviewed on request.

## Appendix materials

**Figure 1:**
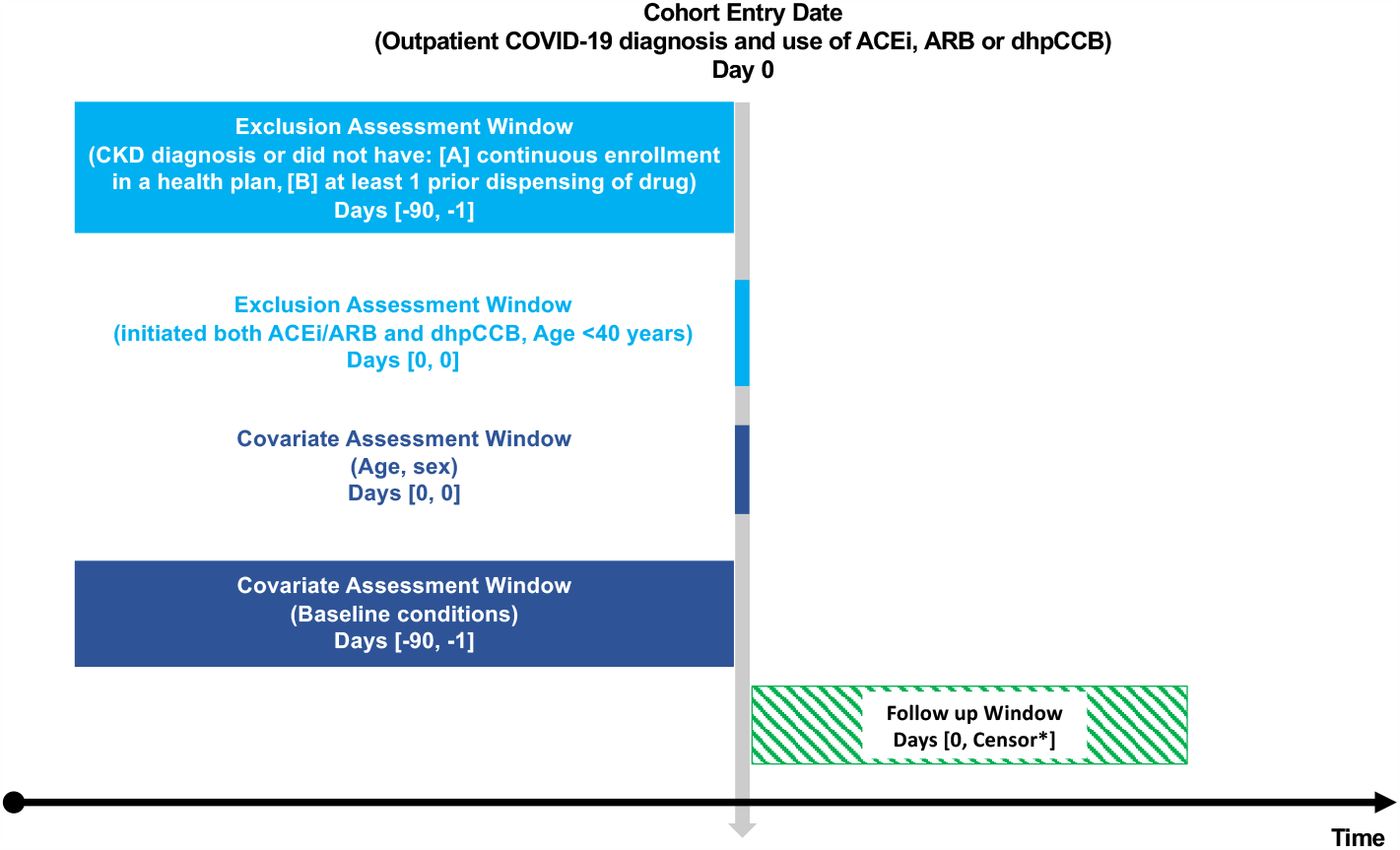
Cohort study design diagram. Abbreviations: ARB, angiotensin receptor blocker; ACEi, angiotensin converting enzyme inhibitor; dhpCCB, dihydropyridine calcium channel blocker; CKD, chronic kidney disease. ^*^Follow-up started the day after cohort entry until the earliest occurrence of a censoring event: outcome, 30 days after cohort entry, death, disenrollment, or end of study period (May 30, 2020).

### Appendix A Coding algorithms for population and exposure

<u>REFERENT: dhpCCB (</u>Excludes: verapamil and diltiazem).

- The occurrence of Pharmacy Claims with the following attributes:
  - Generic Name is any of: { “CLEVIDIPINE BUTYRATE”, “ALISKIREN HEMIFUMARATE/AMLODIPINE/HYDROCHLOROTHIAZIDE”, “MIBEFRADIL DI-HCL”, “ALISKIREN HEMIFUMARATE/AMLODIPINE BESYLATE”, “BEPRIDIL HCL”, “NICARDIPINE IN DEXTROSE, ISO-OSMOTIC”, “NICARDIPINE IN SODIUM CHLORIDE, ISO-OSMOTIC”, “NIMODIPINE”, “ISRADIPINE”, “NICARDIPINE HCL”, “AMLODIPINE BESYLATE/ATORVASTATIN CALCIUM”, “FELODIPINE”, “AMLODIPINE BESYLATE”, “NIFEDIPINE” }
    - CLEVIDIPINE BUTYRATE
    - MIBEFRADIL DI-HCL
    - BEPRIDIL HCL
    - NICARDIPINE IN DEXTROSE, ISO-OSMOTIC
    - NICARDIPINE IN SODIUM CHLORIDE, ISO-OSMOTIC
    - NIMODIPINE
    - ISRADIPINE
    - NICARDIPINE HCL
    - AMLODIPINE BESYLATE/ATORVASTATIN CALCIUM
    - FELODIPINE
    - AMLODIPINE BESYLATE
    - NIFEDIPINE

<u>EXPOSURE: ACEi</u>

- The occurrence of Pharmacy Claims with the following attributes:
  - Generic Name is any of: { “AMLODIPINE BESYLATE/BENAZEPRIL HCL”, “BENAZEPRIL HCL”, “BENAZEPRIL HCL/HYDROCHLOROTHIAZIDE”, “CAPTOPRIL”, “CAPTOPRIL/HYDROCHLOROTHIAZIDE”, “ENALAPRIL MALEATE”, “ENALAPRIL MALEATE/DILTIAZEM MALATE”, “ENALAPRIL MALEATE/FELODIPINE”, “ENALAPRIL MALEATE/HYDROCHLOROTHIAZIDE”, “ENALAPRILAT DIHYDRATE”, “FOSINOPRIL SODIUM”, “FOSINOPRIL SODIUM/HYDROCHLOROTHIAZIDE”, “LISINOPRIL”, “LISINOPRIL/DIETARY SUPPLEMENT,COMB.10”, “LISINOPRIL/HYDROCHLOROTHIAZIDE”, “MOEXIPRIL HCL”, “MOEXIPRIL HCL/HYDROCHLOROTHIAZIDE”, “PERINDOPRIL ARGININE/AMLODIPINE BESYLATE”, “PERINDOPRIL ERBUMINE”, “QUINAPRIL HCL”, “QUINAPRIL HCL/HYDROCHLOROTHIAZIDE”, “RAMIPRIL”, “TRANDOLAPRIL”, “TRANDOLAPRIL/VERAPAMIL HCL” }
    - BENAZEPRIL HCL
    - BENAZEPRIL HCL/HYDROCHLOROTHIAZIDE
    - CAPTOPRIL
    - CAPTOPRIL/HYDROCHLOROTHIAZIDE
    - ENALAPRIL MALEATE
    - ENALAPRIL MALEATE/HYDROCHLOROTHIAZIDE
    - ENALAPRILAT DIHYDRATE
    - FOSINOPRIL SODIUM
    - FOSINOPRIL SODIUM/HYDROCHLOROTHIAZIDE
    - LISINOPRIL
    - LISINOPRIL/DIETARY SUPPLEMENT,COMB.10
    - LISINOPRIL/HYDROCHLOROTHIAZIDE
    - MOEXIPRIL HCL
    - MOEXIPRIL HCL/HYDROCHLOROTHIAZIDE
    - PERINDOPRIL ERBUMINE
    - QUINAPRIL HCL
    - QUINAPRIL HCL/HYDROCHLOROTHIAZIDE
    - RAMIPRIL
    - TRANDOLAPRIL
- The occurrence of Pharmacy Claims with the following attributes:
  - Brand Name is any of: { “ACCUPRIL”, “ACEON”, “ALTACE”, “CAPOTEN”, “LOTENSIN”, “LOTENSIN HCT”, “MAVIK”, “MONOPRIL”, “MONOPRIL HCT”, “PRINIVIL”, “UNIVASC”, “VASOTEC”, “VASOTEC I.V.“, “ZESTRIL” }
    - ACCUPRIL
    - ACEON
    - ALTACE
    - CAPOTEN
    - LOTENSIN
    - LOTENSIN HCT
    - MAVIK
    - MONOPRIL
    - MONOPRIL HCT
    - PRINIVIL
    - UNIVASC
    - VASOTEC
    - VASOTEC I.V.
    - ZESTRIL

<u>EXPOSURE: ARB</u>

- The occurrence of Pharmacy Claims with the following attributes:
  - Generic Name is any of: { “ALISKIREN/VALSARTAN”, “AMLODIPINE BESYLATE/OLMESARTAN MEDOXOMIL”, “AMLODIPINE BESYLATE/VALSARTAN”, “AMLODIPINE BESYLATE/VALSARTAN/HYDROCHLOROTHIAZIDE”, “AZILSARTAN MEDOXOMIL”, “AZILSARTAN MEDOXOMIL/CHLORTHALIDONE”, “CANDESARTAN CILEXETIL”, “CANDESARTAN CILEXETIL/HYDROCHLOROTHIAZIDE”, “EPROSARTAN MESYLATE”, “EPROSARTAN MESYLATE/HYDROCHLOROTHIAZIDE”, “IRBESARTAN”, “IRBESARTAN/HYDROCHLOROTHIAZIDE”, “LOSARTAN POTASSIUM”, “LOSARTAN POTASSIUM/HYDROCHLOROTHIAZIDE”, “NEBIVOLOL HCL/VALSARTAN”, “OLMESARTAN MEDOXOMIL”, “OLMESARTAN MEDOXOMIL/AMLODIPINE BESYLATE/HYDROCHLOROTHIAZIDE”, “OLMESARTAN MEDOXOMIL/HYDROCHLOROTHIAZIDE”, “SACUBITRIL/VALSARTAN”, “TELMISARTAN”, “TELMISARTAN/AMLODIPINE BESYLATE”, “TELMISARTAN/HYDROCHLOROTHIAZIDE”, “VALSARTAN”, “VALSARTAN/HYDROCHLOROTHIAZIDE” }
    - ALISKIREN/VALSARTAN
    - AZILSARTAN MEDOXOMIL
    - AZILSARTAN MEDOXOMIL/CHLORTHALIDONE
    - CANDESARTAN CILEXETIL
    - CANDESARTAN CILEXETIL/HYDROCHLOROTHIAZIDE
    - EPROSARTAN MESYLATE
    - EPROSARTAN MESYLATE/HYDROCHLOROTHIAZIDE
    - IRBESARTAN
    - IRBESARTAN/HYDROCHLOROTHIAZIDE
    - LOSARTAN POTASSIUM
    - LOSARTAN POTASSIUM/HYDROCHLOROTHIAZIDE
    - NEBIVOLOL HCL/VALSARTAN
    - OLMESARTAN MEDOXOMIL
    - OLMESARTAN MEDOXOMIL/HYDROCHLOROTHIAZIDE
    - SACUBITRIL/VALSARTAN
    - TELMISARTAN
    - TELMISARTAN/HYDROCHLOROTHIAZIDE
    - VALSARTAN
    - VALSARTAN/HYDROCHLOROTHIAZIDE
- The occurrence of Pharmacy Claims with the following attributes:
  - Brand Name is any of: { “ATACAND”, “ATACAND HCT”, “AVALIDE”, “AVAPRO”, “BENICAR”, “BENICAR HCT”, “COZAAR”, “DIOVAN”, “DIOVAN HCT”, “EDARBI”, “EXFORGE”, “EXFORGE HCT”, “HYZAAR”, “MICARDIS”, “MICARDIS HCT”, “TEVETEN”, “TEVETEN HCT” }
    - ATACAND
    - ATACAND HCT
    - AVALIDE
    - AVAPRO
    - BENICAR
    - BENICAR HCT
    - COZAAR
    - DIOVAN
    - DIOVAN HCT
    - EDARBI
    - EXFORGE
    - EXFORGE HCT
    - HYZAAR
    - MICARDIS
    - MICARDIS HCT
    - TEVETEN
    - TEVETEN HCT

<u>COVID-19 diagnosis:</u>

- This measure creates a logical OR of events, and identifies all occurrences of one or more specified events:
  - COVID-19, virus identified, outpatient (U07.1)
  - SARS-CoV-2 viral lab test (labs) with positive or presumptive positive result
  - Other coronavirus infection (not U07.1)

<u>COVID-19, virus identified:</u>

- The occurrence of Medical Claims with the following attributes:
  - Diagnosis Code, ICD-10 is any of: { “U07.1” }
    - U07.1 - COVID-19, virus identified

<u>COVID-19 viral lab test, with positive of presumed positive result:</u>

- The occurrence of Lab Tests with the following attributes:
  - Test Ordered Name is any of: { “COVID-19 Nasopharynx”, “COVID-19 Oropharynx”, “COVID-19 Pooled NP/OP”, “SARS CoV 2 RNA(COVID 19), QL NAAT”, “SARS Coronavirus with CoV-2 RNA, Quant RT-PCR”, “SARS-CoV-2 RNA,QL REAL-TIME RT-PCR (COVID-19)“, “SARS-CoV-2 RNA,QL REAL-TIME RT-PCR (COVID-19) - Non-swab”, “COVID-19 Nasal/Nasopharynx”, “SARS CoV 2 RNA, QL Real Time RT PCR”, “SARS CoV 2 RNA, RT-PCR”, “SARS-COV-2 RNA (COVID-19), QUALITATIVE NAAT”, “SARS-CoV-2 RNA, QL, RT PCR (COVID-19) - Swabs”, “SARS-CoV-2 RNA, QL, RT PCR (COVID-19) Swabs”, “COVID-19 Pooled N/NP/OP”, “SARS CORONAVIRUS WITH COV-2 RNA, QUALITATIVE REAL-TIME RTPCR”, “SARS COV 2 RNA, RT PCR”, “SARS CoV 2 RNA(COVID 19), QUALITATIVE NAAT”, “SARS COV W/COV 2 RNA,PCR”, “SARS CoV 2 RNA, QL REAL TIME RT PCR”, “SARS Coronavirus w/CoV 2 RNA,QL Real Time RT PCR”, “SARS-CoV-2 RNA,QL Real-Time RT-PCR (CoVID-19) - Non-swab”, “SARS CORONAVIRUS W/CoV 2 RNA,QL REAL TIME RT PCR”, “SARS-COV-2 RNA, QL REAL-TIME RT-PCR”, “SARS-CoV-2 RNA (COVID-19), Qualitative NAAT”, “SARS COV 2 RNA, QL NAAT”, “SARS Cov 2 RNA, QL NAAT” }
    - COVID-19 Nasopharynx
    - COVID-19 Oropharynx
    - COVID-19 Pooled NP/OP
    - SARS CoV 2 RNA(COVID 19), QL NAAT
    - SARS Coronavirus with CoV-2 RNA, Quant RT-PCR
    - SARS-CoV-2 RNA,QL REAL-TIME RT-PCR (COVID-19)
    - SARS-CoV-2 RNA,QL REAL-TIME RT-PCR (COVID-19) - Non-swab
    - COVID-19 Nasal/Nasopharynx
    - SARS CoV 2 RNA, QL Real Time RT PCR
    - SARS CoV 2 RNA, RT-PCR
    - SARS-COV-2 RNA (COVID-19), QUALITATIVE NAAT
    - SARS-CoV-2 RNA, QL, RT PCR (COVID-19) - Swabs
    - SARS-CoV-2 RNA, QL, RT PCR (COVID-19) Swabs
    - COVID-19 Pooled N/NP/OP
    - SARS CORONAVIRUS WITH COV-2 RNA, QUALITATIVE REAL-TIME RTPCR
    - SARS COV 2 RNA, RT PCR
    - SARS CoV 2 RNA(COVID 19), QUALITATIVE NAAT
    - SARS COV W/COV 2 RNA,PCR
    - SARS CoV 2 RNA, QL REAL TIME RT PCR
    - SARS Coronavirus w/CoV 2 RNA,QL Real Time RT PCR
    - SARS-CoV-2 RNA,QL Real-Time RT-PCR (CoVID-19) - Non-swab
    - SARS CORONAVIRUS W/CoV 2 RNA,QL REAL TIME RT PCR
    - SARS-COV-2 RNA, QL REAL-TIME RT-PCR
    - SARS-CoV-2 RNA (COVID-19), Qualitative NAAT
    - SARS COV 2 RNA, QL NAAT
    - SARS Cov 2 RNA, QL NAAT
  - Result is any of: { “Positive for 2019-nCoV”, “Presumptive Pos. for 2019-nCoV”, “Presumptive Positive 2019-nCoV”, “Presumptive Positive for 2019-nCoV” }
    - Positive for 2019-nCoV
    - Presumptive Pos. for 2019-nCoV
    - Presumptive Positive 2019-nCoV
    - Presumptive Positive for 2019-nCoV
- The occurrence of Lab Tests with the following attributes:
  - Result Name is any of: { “COVID-19 SWB”, “SARS CoV 2 RNA, QL Real Time RT PCR”, “SARS Coronavirus w/CoV 2 RNA,QL Real Time RT PCR”, “COVID-19 Nasopharynx”, “SARS-CoV-2 RNA”, “SARS-CoV-2 RNA,QL REAL-TIME RT-PCR (COVID-19)“, “SARS-CoV-2 RNA,QL REAL-TIME RT-PCR (COVID-19) - Non-swab”, “COVID-19 Nasal/Nasopharynx”, “COVID-19 Oropharynx”, “COVID-19 Pooled N/NP/OP”, “COVID-19 Pooled NP/OP”, “SARS CoV 2 RNA(COVID 19), QL NAAT”, “SARS CoV 2 RNA:“, “SARS CoV 2 RNA”, “SARS-CoV-2 RNA,QL Real-Time RT-PCR (CoVID-19) - Non-swab”, “SARS COV 2 RNA, RT PCR”, “SARS CoV 2 RNA, RT PCR”, “SARS-COV-2 RNA:“, “SARS-CoV-2 RNA:” }
    - COVID-19 SWB
    - SARS CoV 2 RNA, QL Real Time RT PCR
    - SARS Coronavirus w/CoV 2 RNA,QL Real Time RT PCR
    - COVID-19 Nasopharynx
    - SARS-CoV-2 RNA
    - SARS-CoV-2 RNA,QL REAL-TIME RT-PCR (COVID-19)
    - SARS-CoV-2 RNA,QL REAL-TIME RT-PCR (COVID-19) - Non-swab
    - COVID-19 Nasal/Nasopharynx
    - COVID-19 Oropharynx
    - COVID-19 Pooled N/NP/OP
    - COVID-19 Pooled NP/OP
    - SARS CoV 2 RNA(COVID 19), QL NAAT
    - SARS CoV 2 RNA:
    - SARS CoV 2 RNA
    - SARS-CoV-2 RNA,QL Real-Time RT-PCR (CoVID-19) - Non-swab
    - SARS COV 2 RNA, RT PCR
    - SARS CoV 2 RNA, RT PCR
    - SARS-COV-2 RNA:
    - SARS-CoV-2 RNA:
  - Result is any of: { “Presumptive Positive for 2019-nCoV”, “Presumptive Pos. for 2019-nCoV”, “Positive for 2019-nCoV”, “Presumptive Positive 2019-nCoV” }
    - Presumptive Positive for 2019-nCoV
    - Presumptive Pos. for 2019-nCoV
    - Positive for 2019-nCoV
    - Presumptive Positive 2019-nCoV
- The occurrence of Lab Tests with the following attributes:
  - LOINC Code is any of: { “943092”, “945006” }
    - 943092
    - 945006
  - Result is any of: { “Presumptive Positive for 2019-nCoV”, “Presumptive Pos. for 2019-nCoV”, “Positive for 2019-nCoV”, “Presumptive Positive 2019-nCoV” }
    - Presumptive Positive for 2019-nCoV
    - Presumptive Pos. for 2019-nCoV
    - Positive for 2019-nCoV
    - Presumptive Positive 2019-nCoV
- The occurrence of Lab Tests with the following attributes:
  - Procedure Code, HCPCS and CPT is any of: { “U0002”, “87635”, “U0001” }
    - U0002 - Non-CDC laboratory test for SARS-CoV-2/2019-nCoV (COVID-19)
    - 87635 - Infectious agent detection by nucleic acid (DNA or RNA); severe acute respiratory syndrome coronavirus 2 (SARS-CoV-2) (Coronavirus disease [COVID-19]), amplified probe technique
    - U0001 - CDC laboratory test for SARS-CoV-2/2019-nCoV (COVID-19)
  - Result is any of: { “Presumptive Positive for 2019-nCoV”, “Presumptive Pos. for 2019-nCoV”, “Positive for 2019-nCoV”, “Presumptive Positive 2019-nCoV” }
    - Presumptive Positive for 2019-nCoV
    - Presumptive Pos. for 2019-nCoV
    - Positive for 2019-nCoV
    - Presumptive Positive 2019-nCoV

<u>Other coronavirus infection (not U07.1):</u>

- This measure creates a logical OR of events, and identifies all occurrences of one or more specified events:
  - B34.2 - Coronavirus infection, unspecified
  - B97.29 - Other coronavirus as the cause of diseases classified elsewhere
  - B97.21 - SARS-associated coronavirus as the cause of diseases classified elsewhere
  - J12.81 - Pneumonia due to SARS-associated coronavirus

### Appendix B Coding algorithms for outcomes

<u>Hospitalization with COVID-19 diagnosis:</u>

- The occurrence of Medical Claims with the following attributes:
- Diagnosis Code, ICD-10 is any of: { “U07.1” }
  - U07.1 - COVID-19, virus identified
- Inpatient Indicator is any of:
  - Yes
- The occurrence of Medical Claims with the following attributes:
  - Diagnosis Code, ICD-10 is any of: { “U07.1” }
    - U07.1 - COVID-19, virus identified
  - Hospital Service Revenue Code is any of: { “0012”, “0100”, “0110”, “0116”, “0123”, “0125”, “0129”, “0132”, “0144”, “0145”, “0152”, “0154”, “0159”, “0118”, “0121”, “0124”, “0127”, “0130”, “0134”, “0136”, “0139”, “0140”, “0150”, “0158”, “0160”, “0167”, “0987”, “0115”, “0117”, “0119”, “0133”, “0138”, “0141”, “0143”, “0147”, “0148”, “0153”, “0155”, “0164”, “0112”, “0113”, “0114”, “0120”, “0122”, “0126”, “0128”, “0131”, “0135”, “0151”, “0157”, “0169”, “0101”, “0111”, “0142”, “0156” }}

<u>Hospitalization with COVID-19 + the occurrence of ARDS:</u>

This measure was defined as:

- 1.1. The occurrence of Hospitalization with COVID
- 1.2. The occurrence of ARDS, acute respiratory failure, and arrest, starting at least 0 days after the start of 1.1, and starting at most 30 days after the start of 1.1.
- The event of interest in this group is 1.2.

<u>The occurrence of ARDS, acute respiratory failure, and arrest</u>

- The occurrence of Medical Claims with the following attributes:
  - Diagnosis Code, ICD-10 is any of: { “R09.2”, “J80”, “J96.02”, “J96.20”, “J96.21”, “J96.22”, “J96.00”, “J96.2”, “J96.0”, “J96.01” }
    - R09.2 - Respiratory arrest
    - J80 - Acute respiratory distress syndrome
    - J96.02 - Acute respiratory failure with hypercapnia
    - J96.20 - Acute and chronic respiratory failure, unspecified whether with hypoxia or hypercapnia
    - J96.21 - Acute and chronic respiratory failure with hypoxia
    - J96.22 - Acute and chronic respiratory failure with hypercapnia
    - J96.00 - Acute respiratory failure, unspecified whether with hypoxia or hypercapnia
    - J96.2 - Acute and chronic respiratory failure
    - J96.0 - Acute respiratory failure
    - J96.01 - Acute respiratory failure with hypoxia

<u>Hospitalization for COVID-19 + the occurrence of Acute organ dysfunction or failure:</u>

This measure was defined as:

- 1.1. The occurrence of Hospitalization with COVID
- 1.2. The occurrence of Inpatient hospitalization with acute organ dysfunction or failure, starting at least 0 days after the start of 1.1, and starting at most 30 days after the start of 1.1.
- The event of interest in this group is 1.2.

<u>The occurrence of Acute organ dysfunction or failure:</u>

- The occurrence of Medical Claims with the following attributes:
  - Diagnosis Code, ICD-10 is any of: { “G62.81”, “N17.9”, “G93.41”, “J96.0”, “J96.01”, “K72.01”, “N17”, “N17.1”, “D65”, “K72.0”, “K72.00”, “N17.2”, “J96.00”, “J96.02”, “N17.8”, “G72.81”, “N17.0” }}
    - G62.81 - Critical illness polyneuropathy
    - N17.9 - Acute kidney failure, unspecified
    - G93.41 - Metabolic encephalopathy
    - J96.0 - Acute respiratory failure
    - J96.01 - Acute respiratory failure with hypoxia
    - K72.01 - Acute and subacute hepatic failure with coma
    - N17 - Acute kidney failure
    - N17.1 - Acute kidney failure with acute cortical necrosis
    - D65 - Disseminated intravascular coagulation [defibrination syndrome]
    - K72.0 - Acute and subacute hepatic failure
    - K72.00 - Acute and subacute hepatic failure without coma
    - N17.2 - Acute kidney failure with medullary necrosis
    - J96.00 - Acute respiratory failure, unspecified whether with hypoxia or hypercapnia
    - J96.02 - Acute respiratory failure with hypercapnia
    - N17.8 - Other acute kidney failure
    - G72.81 - Critical illness myopathy
    - N17.0 - Acute kidney failure with tubular necrosis
  - Inpatient Indicator is any of:
    - Yes
- The occurrence of Medical Claims with the following attributes:
  - Diagnosis Code, ICD-10 is any of: { “G62.81”, “N17.9”, “G93.41”, “J96.0”, “J96.01”, “K72.01”, “N17”, “N17.1”, “D65”, “K72.0”, “K72.00”, “N17.2”, “J96.00”, “J96.02”, “N17.8”, “G72.81”, “N17.0” }
  - Suspected Inpatient Indicator is any of:
    - Yes

<u>Hospitalization with COVID-19 + the occurrence of Sepsis:</u>

This measure was defined as:

- 1.1. The occurrence of Hospitalization with COVID
- 1.2. The occurrence of Occurrence of sepsis inpatient, starting at least 0 days after the start of 1.1, and starting at most 30 days after the start of 1.1.
- The event of interest in this group is 1.2.

<u>The occurrence of sepsis, inpatient:</u>

- The occurrence of Medical Claims with the following attributes:
  - Diagnosis Code, ICD-10 is any of: { “A02.1”, “A26.7”, “A40”, “A40.1”, “A41.01”, “A41.1”, “A41.59”, “B96.2”, “P35.2”, “A22.7”, “A28.0”, “A32.7”, “A39.4”, “A40.0”, “A40.3”, “A41.52”, “B37.7”, “I76”, “N39.0”, “R65.20”, “A20.7”, “A41”, “A41.02”, “A41.4”, “A41.51”, “A41.89”, “A41.9”, “B00.7”, “B95.61”, “B95.62”, “P36.0”, “P36.2”, “P36.8”, “R65.2”, “A28.2”, “A39.2”, “A41.0”, “A41.2”, “A41.5”, “A41.53”, “A41.8”, “A42.7”, “J44.0”, “P36.1”, “P36.3”, “P36.4”, “P36.9”, “P37.2”, “P37.5”, “A03.9”, “A04.7”, “A21.7”, “A23.9”, “A24.1”, “A39.3”, “A40.8”, “A40.9”, “A41.3”, “A41.50”, “A41.81”, “A54.86”, “B95.4”, “B95.6”, “J18.9”, “P36.5”, “R65.21” }
    - A02.1 - Salmonella sepsis
    - A26.7 - Erysipelothrix sepsis
    - A40 - Streptococcal sepsis
    - A40.1 - Sepsis due to streptococcus, group B
    - A41.01 - Sepsis due to Methicillin susceptible Staphylococcus aureus
    - A41.1 - Sepsis due to other specified staphylococcus
    - A41.59 - Other Gram-negative sepsis
    - B96.2 - Escherichia coli [E. coli] as the cause of diseases classified elsewhere
    - P35.2 - Congenital herpesviral [herpes simplex] infection
    - A22.7 - Anthrax sepsis
    - A28.0 - Pasteurellosis
    - A32.7 - Listerial sepsis
    - A39.4 - Meningococcemia, unspecified
    - A40.0 - Sepsis due to streptococcus, group A
    - A40.3 - Sepsis due to Streptococcus pneumoniae
    - A41.52 - Sepsis due to Pseudomonas
    - B37.7 - Candidal sepsis
    - I76 - Septic arterial embolism
    - N39.0 - Urinary tract infection, site not specified
    - R65.20 - Severe sepsis without septic shock
    - A20.7 - Septicemic plague
    - A41 - Other sepsis
    - A41.02 - Sepsis due to Methicillin resistant Staphylococcus aureus
    - A41.4 - Sepsis due to anaerobes
    - A41.51 - Sepsis due to Escherichia coli [E. coli]
    - A41.89 - Other specified sepsis
    - A41.9 - Sepsis, unspecified organism
    - B00.7 - Disseminated herpesviral disease
    - B95.61 - Methicillin susceptible Staphylococcus aureus infection as the cause of diseases classified elsewhere
    - B95.62 - Methicillin resistant Staphylococcus aureus infection as the cause of diseases classified elsewhere
    - P36.0 - Sepsis of newborn due to streptococcus, group B
    - P36.2 - Sepsis of newborn due to Staphylococcus aureus
    - P36.8 - Other bacterial sepsis of newborn
    - R65.2 - Severe sepsis
    - A28.2 - Extraintestinal yersiniosis
    - A39.2 - Acute meningococcemia
    - A41.0 - Sepsis due to Staphylococcus aureus
    - A41.2 - Sepsis due to unspecified staphylococcus
    - A41.5 - Sepsis due to other Gram-negative organisms
    - A41.53 - Sepsis due to Serratia
    - A41.8 - Other specified sepsis
    - A42.7 - Actinomycotic sepsis
    - J44.0 - Chronic obstructive pulmonary disease with acute lower respiratory infection
    - P36.1 - Sepsis of newborn due to other and unspecified streptococci
    - P36.3 - Sepsis of newborn due to other and unspecified staphylococci
    - P36.4 - Sepsis of newborn due to Escherichia coli
    - P36.9 - Bacterial sepsis of newborn, unspecified
    - P37.2 - Neonatal (disseminated) listeriosis
    - P37.5 - Neonatal candidiasis
    - A03.9 - Shigellosis, unspecified
    - A04.7 - Enterocolitis due to Clostridium difficile
    - A21.7 - Generalized tularemia
    - A23.9 - Brucellosis, unspecified
    - A24.1 - Acute and fulminating melioidosis
    - A39.3 - Chronic meningococcemia
    - A40.8 - Other streptococcal sepsis
    - A40.9 - Streptococcal sepsis, unspecified
    - A41.3 - Sepsis due to Hemophilus influenzae
    - A41.50 - Gram-negative sepsis, unspecified
    - A41.81 - Sepsis due to Enterococcus
    - A54.86 - Gonococcal sepsis
    - B95.4 - Other streptococcus as the cause of diseases classified elsewhere
    - B95.6 - Staphylococcus aureus as the cause of diseases classified elsewhere
    - J18.9 - Pneumonia, unspecified organism
    - P36.5 - Sepsis of newborn due to anaerobes
    - R65.21 - Severe sepsis with septic shock
  - Inpatient Indicator is any of:
    - Yes
- The occurrence of Medical Claims with the following attributes:
  - Diagnosis Code, ICD-10 is any of: { “A02.1”, “A26.7”, “A40”, “A40.1”, “A41.01”, “A41.1”, “A41.59”, “B96.2”, “P35.2”, “A22.7”, “A28.0”, “A32.7”, “A39.4”, “A40.0”, “A40.3”, “A41.52”, “B37.7”, “I76”, “N39.0”, “R65.20”, “A20.7”, “A41”, “A41.02”, “A41.4”, “A41.51”, “A41.89”, “A41.9”, “B00.7”, “B95.61”, “B95.62”, “P36.0”, “P36.2”, “P36.8”, “R65.2”, “A28.2”, “A39.2”, “A41.0”, “A41.2”, “A41.5”, “A41.53”, “A41.8”, “A42.7”, “J44.0”, “P36.1”, “P36.3”, “P36.4”, “P36.9”, “P37.2”, “P37.5”, “A03.9”, “A04.7”, “A21.7”, “A23.9”, “A24.1”, “A39.3”, “A40.8”, “A40.9”, “A41.3”, “A41.50”, “A41.81”, “A54.86”, “B95.4”, “B95.6”, “J18.9”, “P36.5”, “R65.21” }
  - Suspected Inpatient Indicator is any of:
    - Yes
- The occurrence of Medical Claims with the following attributes:
  - Diagnosis Code, ICD-10 is any of: { “A02.1”, “A26.7”, “A40”, “A40.1”, “A41.01”, “A41.1”, “A41.59”, “B96.2”, “P35.2”, “A22.7”, “A28.0”, “A32.7”, “A39.4”, “A40.0”, “A40.3”, “A41.52”, “B37.7”, “I76”, “N39.0”, “R65.20”, “A20.7”, “A41”, “A41.02”, “A41.4”, “A41.51”, “A41.89”, “A41.9”, “B00.7”, “B95.61”, “B95.62”, “P36.0”, “P36.2”, “P36.8”, “R65.2”, “A28.2”, “A39.2”, “A41.0”, “A41.2”, “A41.5”, “A41.53”, “A41.8”, “A42.7”, “J44.0”, “P36.1”, “P36.3”, “P36.4”, “P36.9”, “P37.2”, “P37.5”, “A03.9”, “A04.7”, “A21.7”, “A23.9”, “A24.1”, “A39.3”, “A40.8”, “A40.9”, “A41.3”, “A41.50”, “A41.81”, “A54.86”, “B95.4”, “B95.6”, “J18.9”, “P36.5”, “R65.21” }
  - Hospital Service Revenue Code is any of: { “0114”, “0115”, “0129”, “0142”, “0143”, “0144”, “0151”, “0156”, “0987”, “0100”, “0101”, “0118”, “0122”, “0127”, “0133”, “0152”, “0160”, “0117”, “0120”, “0130”, “0136”, “0140”, “0145”, “0150”, “0155”, “0158”, “0169”, “0012”, “0111”, “0113”, “0116”, “0119”, “0121”, “0125”, “0139”, “0141”, “0147”, “0148”, “0154”, “0157”, “0164”, “0167”, “0110”, “0112”, “0123”, “0124”, “0126”, “0128”, “0131”, “0132”, “0134”, “0135”, “0138”, “0153”, “0159” }

<u>Hospitalization with COVID-19 + the occurrence of respiratory intubation and mechanical ventilation (procedure and diagnosis codes):</u>

This measure was defined as:

- 1.1. The occurrence of Hospitalization with COVID
- 1.2. The occurrence of Respiratory intubation and mechanical ventilation (procedure and diagnosis codes), starting at least 0 days after the start of 1.1, and starting at most 30 days after the start of 1.1.
- The event of interest in this group is 1.2.

<u>The occurrence of respiratory intubation and mechanical ventilation (procedure and diagnosis codes):</u>

- The occurrence of Medical Claims with the following attributes:
  - Procedure Code, HCPCS and CPT is any of: { “31500”, “G8569”, “43753”, “94002”, “94657”, “94003”, “94004”, “94656” }
    - 31500 - Intubation, endotracheal, emergency procedure
    - G8569 - PROLONGED POSTOPERATIVE INTUBATION (> 24 HRS) REQUIRED
    - 43753 - Gastric intubation and aspiration(s) therapeutic, necessitating physician’s skill (eg, for gastrointestinal hemorrhage), including lavage if performed
    - 94002 - Ventilation assist and management, initiation of pressure or volume preset ventilators for assisted or controlled breathing; hospital inpatient/observation, initial day
    - 94657 - Ventilation assist and management, initiation of pressure or volume preset ventilators for assisted or controlled breathing; subsequent days
    - 94003 - Ventilation assist and management, initiation of pressure or volume preset ventilators for assisted or controlled breathing; hospital inpatient/observation, each subsequent day
    - 94004 - Ventilation assist and management, initiation of pressure or volume preset ventilators for assisted or controlled breathing; nursing facility, per day
    - 94656 - Ventilation assist and management, initiation of pressure or volume preset ventilators for assisted or controlled breathing; first day
- The occurrence of Medical Claims with the following attributes:
  - Diagnosis Code, ICD-10 is any of: { “Z99.1”, “Z99.11”, “Z99.12”, “J95.851”, “J95.85”, “J95.859”, “T88.4”, “T88.4XXA”, “T88.4XXS” }
    - Z99.1 - Dependence on respirator
    - Z99.11 - Dependence on respirator [ventilator] status
    - Z99.12 - Encounter for respirator [ventilator] dependence during power failure
    - J95.851 - Ventilator associated pneumonia
    - J95.85 - Complication of respirator [ventilator]
    - J95.859 - Other complication of respirator [ventilator]
    - T88.4 - Failed or difficult intubation
    - T88.4XXA - Failed or difficult intubation, initial encounter
    - T88.4XXS - Failed or difficult intubation, sequela
- The occurrence of Medical Claims with the following attributes:
  - Procedure Code, ICD-10 is any of: { “09HN8BZ”, “0BH18EZ”, “0DH57BZ”, “0WHQ73Z”, “0WHQ7YZ”, “5A09457”, “09HN7BZ”, “5A09557”, “0CHY7BZ”, “5A09357”, “5A1945Z”, “5A1955Z”, “0BH17EZ”, “0BH13EZ”, “0CHY8BZ”, “0DH58BZ”, “5A1935Z” }
    - 09HN8BZ - Insertion of Airway into Nasopharynx, Via Natural or Artificial Opening Endoscopic
    - 0BH18EZ - Insertion of Endotracheal Airway into Trachea, Via Natural or Artificial Opening Endoscopic
    - 0DH57BZ - Insertion of Airway into Esophagus, Via Natural or Artificial Opening
    - 0WHQ73Z - Insertion of Infusion Device into Respiratory Tract, Via Natural or Artificial Opening
    - 0WHQ7YZ - Insertion of Other Device into Respiratory Tract, Via Natural or Artificial Opening
    - 5A09457 - Assistance with Respiratory Ventilation, 24-96 Consecutive Hours, Continuous Positive Airway Pressure
    - 09HN7BZ - Insertion of Airway into Nasopharynx, Via Natural or Artificial Opening
    - 5A09557 - Assistance with Respiratory Ventilation, Greater than 96 Consecutive Hours, Continuous Positive Airway Pressure
    - 0CHY7BZ - Insertion of Airway into Mouth and Throat, Via Natural or Artificial Opening
    - 5A09357 - Assistance with Respiratory Ventilation, Less than 24 Consecutive Hours, Continuous Positive Airway Pressure
    - 5A1945Z - Respiratory Ventilation, 24-96 Consecutive Hours
    - 5A1955Z - Respiratory Ventilation, Greater than 96 Consecutive Hours
    - 0BH17EZ - Insertion of Endotracheal Airway into Trachea, Via Natural or Artificial Opening
    - 0BH13EZ - Insertion of Endotracheal Airway into Trachea, Percutaneous Approach
    - 0CHY8BZ - Insertion of Airway into Mouth and Throat, Via Natural or Artificial Opening Endoscopic
    - 0DH58BZ - Insertion of Airway into Esophagus, Via Natural or Artificial Opening Endoscopic
    - 5A1935Z - Respiratory Ventilation, Less than 24 Consecutive Hours

